# Mediators between body mass index and atrial fibrillation: a Mendelian Randomization study

**DOI:** 10.1101/2023.05.16.23290070

**Authors:** Ziting Gao, Jun Xiao, Wuqing Huang

## Abstract

**Background:** Although obesity is a recognized risk factor of atrial fibrillation (AF), the mechanisms are not fully understood. Thus, we aimed to identify the potential mediators between body mass index (BMI) and AF.

**Methods:** We conducted a two-sample Mendelian randomization (MR) analysis using publicly available summary-level data from genome-wide association studies. Univariable MR analyses were applied to identify potential mediators, and then the multivariable MR analyses were conducted to explore the mediated roles of circulating cytokines, metabolic markers and comorbidities in the association between BMI and AF.

**Results:** This MR study found a significant causal association between BMI and AF (OR=1.41, 95%CI=1.33-1.50; P<0.001), which was attenuated to 1.21 (95%CI=1.03-1.43) after being adjusted for leptin, in which 48.78% excess risk was mediated. After further adjustment for leptin and sleep apnoea or coronary heart disease, the association was attenuated to null (adjusted for leptin and sleep apnoea: OR=1.05, 95%CI=0.85-1.30; adjusted for leptin and coronary heart disease: OR=1.08, 95%CI=0.90-1.30), resulting in 87.80% and 80.49% excess risk being mediated, respectively.

**Conclusion:** These results identified an important mediated role of leptin, particularly for individuals with sleep apnoea or coronary heart disease, providing some clues for the underlying mechanisms behind the impact of obesity on AF risk.

**Funding:** Natural Science Foundation of Fujian Province (grant no. 2022J01706) and the Start-up Fund for high-level talents of Fujian Medical University (XRCZX2021026).

## Introduction

Atrial fibrillation (AF) is the most common type of arrhythmia and associated with significant elevated risk of morbidity and mortality, resulting in major public health burden and referred to as a cardiovascular epidemic of the 21st century. The incidence is continuously increasing, the Framingham Heart Study has reported a threefold increase in the incidence of AF during the past 50 years.^1^ AF is closely related to a range of classic cardiovascular risk factors, including modifiable lifestyle factors (eg, obesity, hypertension, diabetes, coronary artery disease, hyperthyroidism, kidney disease),and non-modifiable factors (eg, age, ethnic).^2^ Although a number of risk factors have been linked to a higher risk of AF, the etiology has not been clarified yet.

Obesity is a well-established risk factor for AF.^3-8^ ARIC (Atherosclerosis Risk In Communities) study reported that nearly 20% of atrial fibrillation cases can be attributed to overweight or obesity.^9^ However, the underlying pathways behind obesity and AF remained unclear. Several mechanisms have been proposed to explain this link.^2^ For example, obesity always co-exists with multiple metabolic disorders, such as dyslipidemia, hypertension and diabetes, all of which have ever been reported to increase the risk of AF. Prior epidemiological studies found that obese adults were predisposed to some comorbidities, such as coronary heart disease, obstructive sleep apnoea, hyperthyroidism and so on, besides, complex links were reported between AF and these comorbidities, indicating a potential mediated role of comorbidities between obesity and AF. Furthermore, obesity is related to the circulating level of adipocyte-derived cytokines (i.e., adipokines) as well as inflammatory cytokines. Circulating leptin (the marker of leptin resistance) and C reactive protein (CRP) (an important marker of inflammation) are always correlated, increased level of both is common in obesity and independently associated with risk of multiple cardiovascular diseases, including AF.^10,11^

Given that obesity is becoming an epidemic with an estimation of one third of the population globally being overweight or obese, targeting obesity and its pathways is a promising strategy to largely reduce the incidence of AF.^12^ While obesity is a complex condition, evidence of “obesity paradox” is emerging for cardiovascular diseases, for example, some studies found a more favorable prognosis in AF patients with a higher body mass index (BMI).^13^ Thus, it is important to fully understand the possible pathways in the association of BMI and AF. There are a paucity of studies exploring the potential mediators between obesity and AF, which is challenging to be studied in traditional observational epidemiological studies due to the inherent problems of unmeasurable confounding factors and reverse causality. Mendelian randomization (MR) approach complements traditional epidemiological methods as it contributes to overcoming these flaws by using genetic variants as instrumental variables.^14^ Therefore, in this study, we aimed to extensively explore the potential mediators and identify the dominating mediators in the relationship between BMI and AF via multivariable MR study.

## Methods

### Study design

We used publicly available summary-level data from Genome-Wide Association Studies (GWAS) (**Supplementary Table 1**) to conduct two-sample MR study. MR uses genetic variants strongly correlated with exposure factors as instrumental variables (IVs) to estimate causal effects between exposure and outcomes.^14^ All publicly available studies used in this research had been approved by the relevant institutional review boards and participants had provided informed consents.

According to findings from previous studies regarding risk factors of AF, we categorized the potential mediators into three classes in the present study, including circulating cytokines, metabolic markers and comorbidities. Circulating cytokines include leptin, adiponectin, resistin and CRP; Metabolic markers include low-density lipoprotein cholesterol (LDL-C), high-density lipoprotein cholesterol (HDL-C), total cholesterol (TC), triglycerides (TG), fasting blood glucose, systolic blood pressure and diastolic blood pressure; Obesity-related comorbidities include sleep apnoea, coronary heart disease, stroke, chronic kidney disease and hyperthyroidism. The univariable MR analyses were first applied to investigate the total effect of BMI on AF, and to evaluate the causal relationship between BMI and potential mediators as well as between potential mediators and AF to identify the significant mediators. Next, the multivariable MR analyses were conducted to assess the mediated effects of these significant mediators between BMI and AF.

### Data sources

The GWAS summary-level data of BMI was obtained from the Genetic Investigation of ANthropometric Traits (GIANT) consortium, which is a meta-analysis of genome-wide association studies to investigate the genetic associations with height and body mass index in approximately 700,000 individuals of European ancestry.^15^ For atrial fibrillation, we used the latest meta-analysis of GWAS, which compared a total of 60,620 individuals with atrial fibrillation with 970,216 control individuals of European ancestry from six contributing studies, including the Nord-Trøndelag Health Study, the Diabetes Epidemiology: Collaborative analysis of Diagnostic criteria in Europe study, the Michigan Genomics Initiative, DiscovEHR, UK Biobank, and the Atrial Fibrillation Genetics Consortium.^16^ The GWAS summary-level data of leptin derived from the study covering 56,802 samples and 231,001 SNPs.^17^ For other potential mediators, we used GWAS summary data from CARDIoGRAMplusC4D for coronary heart disease, ISGC Consortium for stroke, FinnGen Consortium for sleep apnoea, ADIPOGen Consortium for adiponectin, Global Lipids Genetics Consortium for lipids, International Consortium of Blood Pressure for blood pressure, and UK biobank for hyperthyroidism. Details of data resources are presented in the **Supplementary Table 1**.

### Genetic instruments selection

We selected all single nucleotide polymorphisms (SNPs) significantly related to BMI or each mediator from corresponding GWAS summary-level data as instrumental variables to proxy each exposure, the statistical significance was set at genome-wide significance levels (P < 5 * 10^−8^). SNPs in linkage disequilibrium were excluded, which was estimated by r^2^ at the cutoff value of 0.001 within 10,000 kb window using European reference panel from 1000 Genomes Project.

### Statistical analyses

#### Primary analyses

Two-sample MR analyses were performed by using “TwoSampleMR” R package. In this study, inverse-variance weighted mendelian randomization (IVW-MR) method was applied to obtain the estimates of the causal associations between exposure and outcomes, which is the most widely used approach. The univariable MR analyses were first conducted to evaluate the causal relationship between BMI and AF. Next, we further applied the univariable MR analyses to identify the potential mediated pathways between BMI and AF by screening out the mediators which had causal associations with both BMI and AF. At last, the mediation effect of these identified mediators between BMI and AF was assessed by including these mediators one by one in each multivariable MR model. To further investigate the mediated role of these factors, we calculated the percentage of excess risk mediated (PERM) with odds ratio (OR) as the formula below:

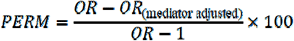

#### Sensitivity analyses

Several sensitivity analyses were performed to assess the robustness of the observed results. The Cochrane Q test was used to test for the heterogeneity of instrumental variables, in which p<0.05 indicates the presence of heterogeneity. The MR-Egger regression intercept and the MR-PRESSO global test was used to assess the horizontal pleiotropy of instrumental variables. Horizontal pleiotropy refers to the fact that SNPs used as instrumental variables can influence the outcome by means other than exposure. In MR-Egger regression, the intercept indicates horizontal pleiotropy; the closer the value is to 0, the less likely it is that the instrumental variable is horizontally pleiotropic, with p<0.05 suggesting possible horizontal pleiotropy. In MR-PRESSO analysis, global test can be used to test for the presence of horizontal pleiotropy where P<0.05 indicates the presence of horizontal pleiotropy. If the horizontal pleiotropy was present, we further identified the horizontal pleiotropic outliers in MR-PRESSO analysis and applied distortion test to test the difference in the estimates before and after outlier removal, where p>0.05 indicates no statistically significant difference. To further identify the SNP primarily affected by pleiotropy, leave-one-out sensitivity analysis was conducted. This involved removing one SNP at a time and calculating the meta-effect of the remaining SNPs. The analysis observed whether there were any changes in the results after the removal of one SNP. SNPs were then rejected one by one to determine whether they significantly affected the results. R software (version 4.1.0) was used to perform all analyses in this study.

## Results

### Univariable MR study for the association of BMI with AF and potential mediators

As shown in **Figure 1** and **Supplementary Table 2**, the univariable MR analysis showed a significant causal association between BMI and AF (OR=1.41, 95%CI=1.33-1.50; P<0.001). For the association with potential mediators, we found that higher level of genetically-predicted BMI was significantly associated with a lower level of systolic blood pressure (OR= 0.58, 95%CI=0.35-0.96; P=0.032), diastolic blood pressure (OR=0.62, 95%CI=0.46-0.84; P=0.002) and with a higher level of leptin (OR=1.86, 95%CI=1.68-2.05; P<0.001), resistin (OR=1.09, 95%CI=1.02-2.16; P=0.008),CRP (OR=1.43, 95%CI=1.37-1.49;P<0.001) and a higher incidence of sleep apnoea (OR=2.13, 95%CI=1.93-2.35; P<0.001), coronary heart disease (OR=1.54, 95%CI=1.43-1.66; P<0.001), stroke (OR=1.22, 95%CI=1.15-1.30; P<0.001), and CKD (OR=1.27, 95%CI=1.16-1.40; P<0.001).

**Figure 1:**
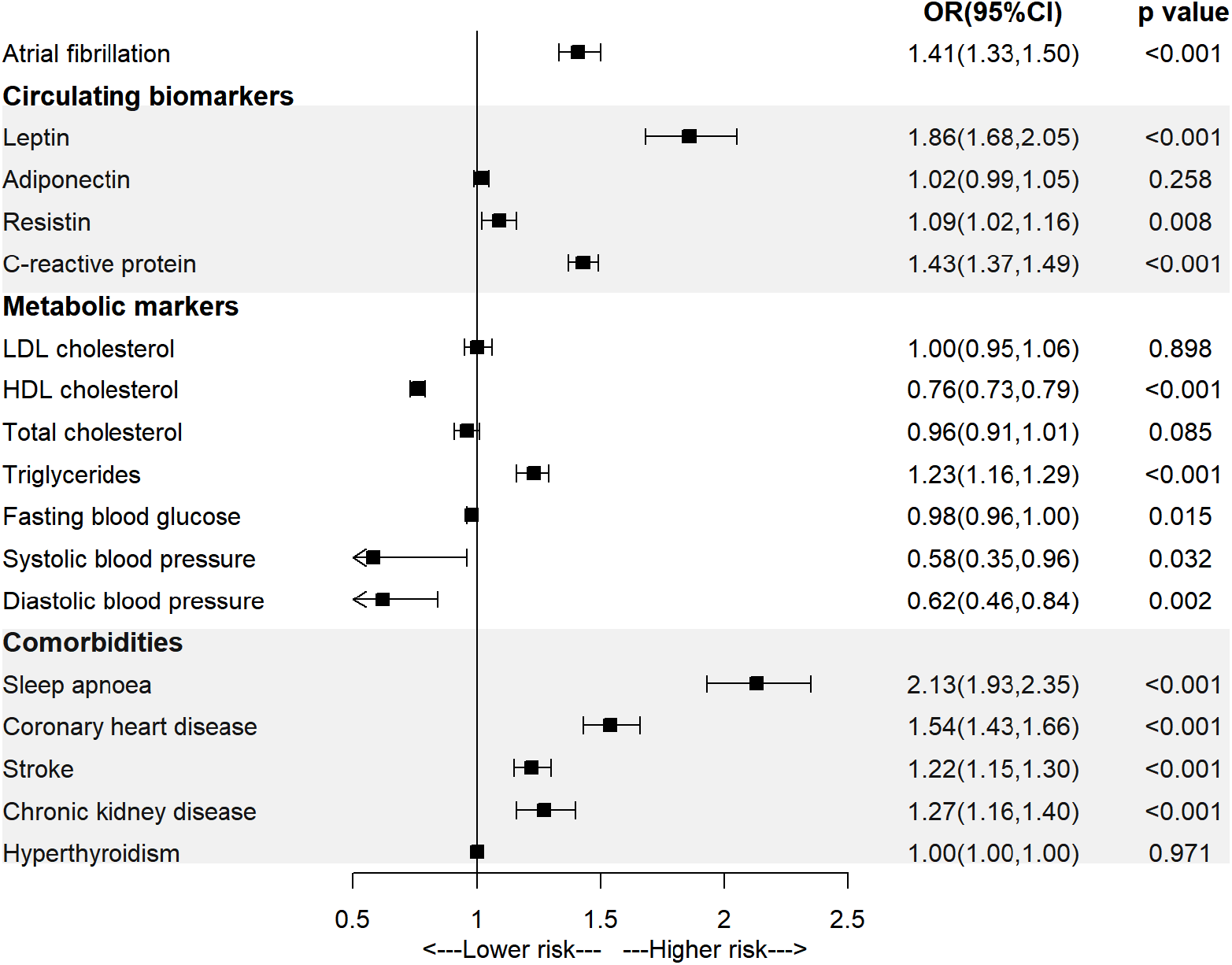
Association of body mass index with atrial fibrillation and mediators in the univariable MR analysis.

### Univariable MR study for the association of potential mediators with AF

As shown in **Figure 2** and **Supplementary Table 3**, the univariable MR analysis observed significant associations between genetically-predicted higher level of leptin (OR=1.31, 95%CI=1.10-1.55; P=0.002), higher level of systolic blood pressure (OR=1.02, 95%CI=1.01-1.02; P<0.001), higher level of diastolic blood pressure (OR=1.03, 95%CI=1.02-1.04; P<0.001), diagnosis of sleep apnoea (OR=1.18, 95%CI=1.07-1.30; P=0.001) or coronary heart disease (OR=1.11, 95%CI=1.05-1.17; P<0.001) and higher incidence of AF.

**Figure 2:**
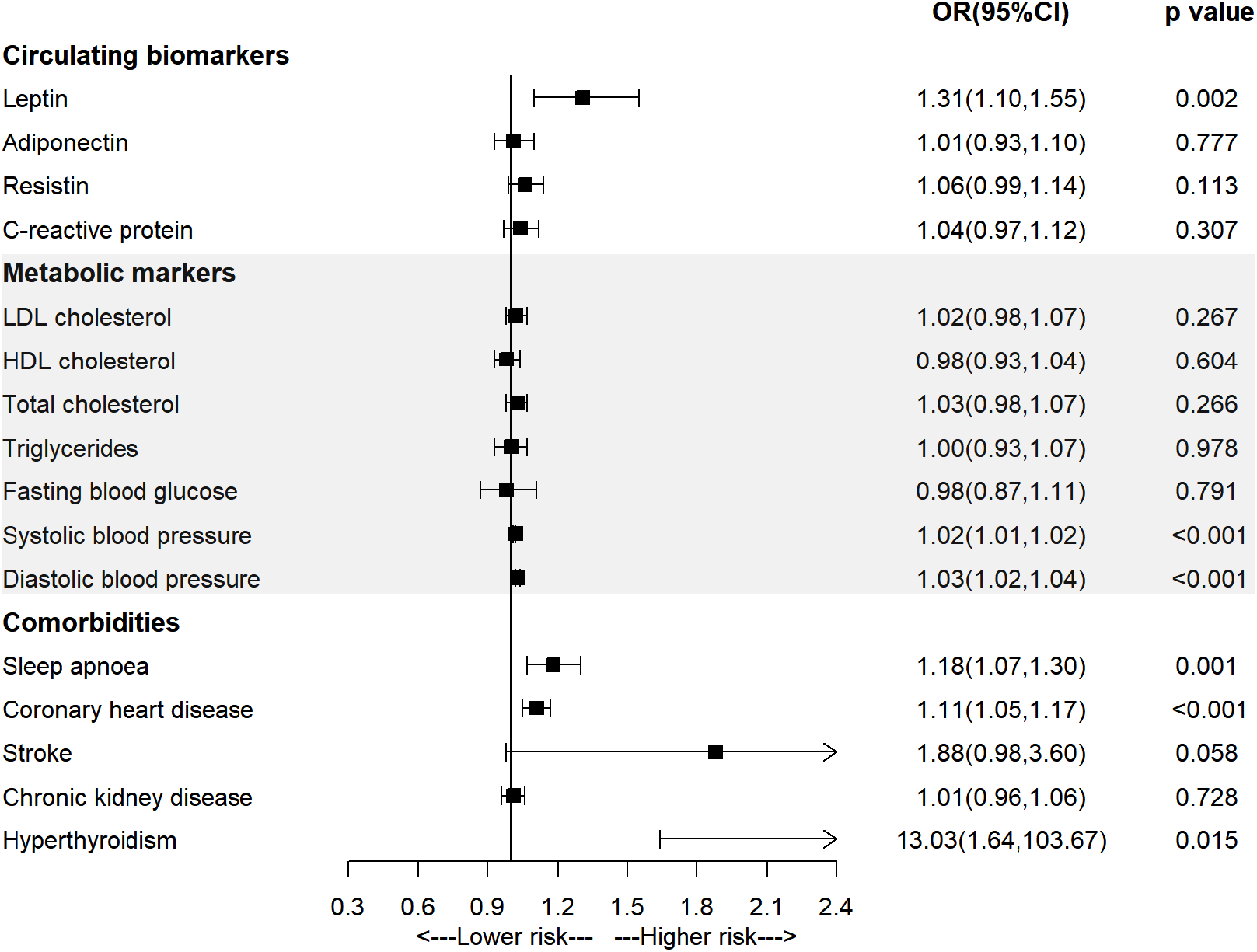
Association of mediators with the risk of atrial fibrillation in the univariable MR analysis.

### Multivariable MR study for the association of BMI with AF

Univariable MR studies identified several factors related to both BMI and AF, including leptin, systolic blood pressure, diastolic blood pressure, sleep apnoea and coronary heart disease, the multivariable MR analyses were performed to assess the mediation effect of these factors. As shown in **Figure 3** and **Supplementary Table 4**, the observed association between BMI and AF was attenuated from 1.41 (95%CI=1.33-1.50) to 1.21 (95%CI=1.03-1.43) after being adjusted for leptin, in which 48.78% excess risk was mediated. After being adjusted for sleep apnoea or coronary heart disease, the association was attenuated to 1.32(95%CI=1.23-1.43) or 1.33(95%CI=1.26-1.42), respectively. Furthermore, the association between BMI and AF was attenuated to null when being adjusted for leptin and each comorbidity (adjusted for leptin and sleep apnoea: OR = 1.05, 95%CI= 0.85-1.30; adjusted for leptin and coronary heart disease: OR = 1.08, 95%CI= 0.90-1.30), resulting in 87.80% and 80.49% excess risk being mediated, respectively. The observed association between BMI and AF did not change significantly after being adjusted for systolic blood pressure, diastolic blood pressure.

**Figure 3:**
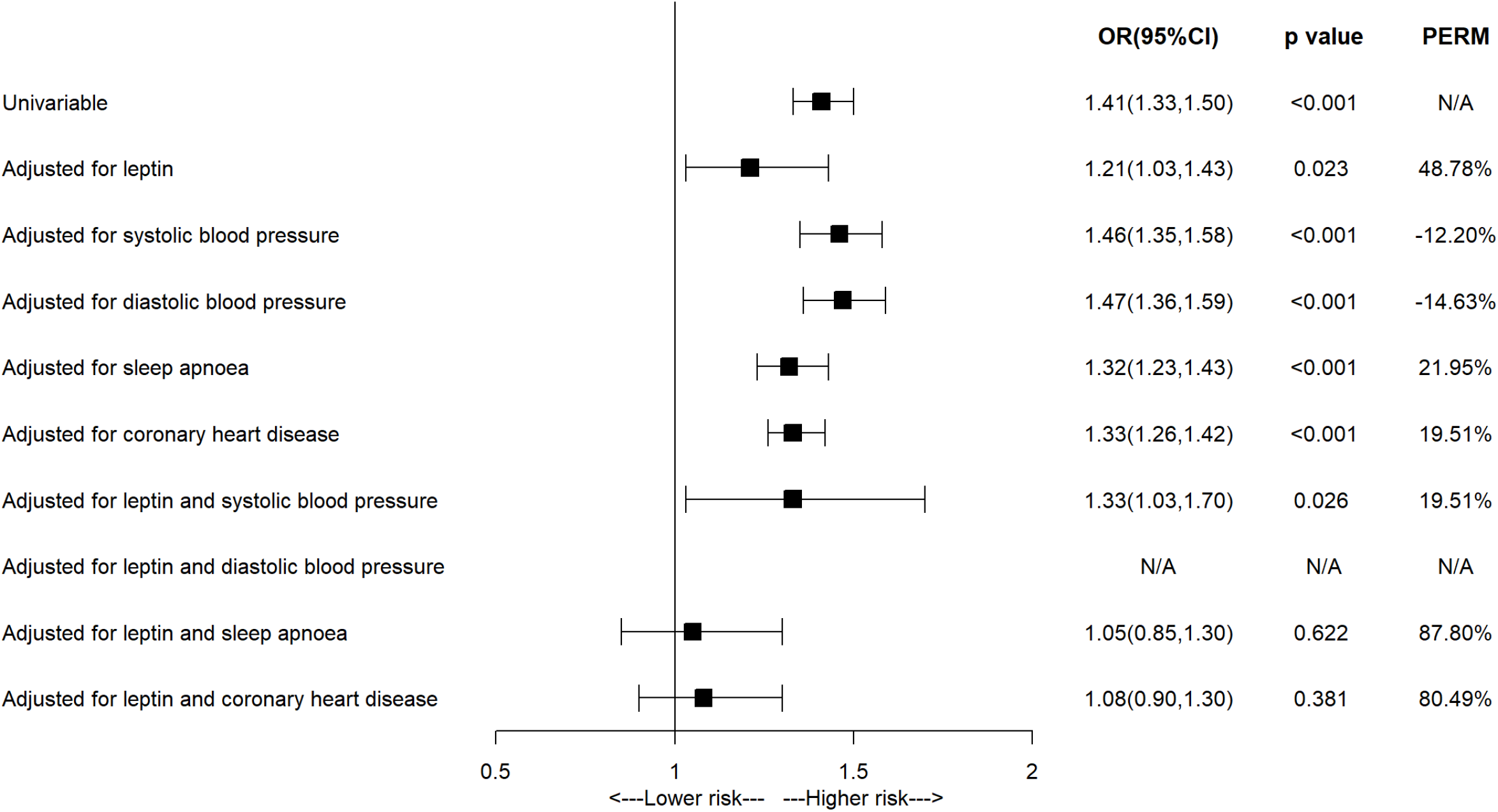
Association of body mass index with the risk of atrial fibrillation in the multivariate MR analysis.

### Sensitivity analyses

The results of sensitivity analyses are present in **Supplementary files**. The scatter plots of the main results showed similar results across different MR methods, suggesting the robustness of the main results **(Supplementary Figure 1)**. The Cochrane Q test showed that potential heterogeneity was present in most analyses except for BMI on CKD, leptin on AF, adiponectin on AF, sleep apnoea on AF, CKD on AF and hyperthyroidism on AF. While leave-one-out analyses suggested that heterogeneity played a small role in the main results **(Supplementary Figure 2-6)**. The results of MR-PRESSO analysis indicated the presence of horizontal pleiotropy in most analyses (P<0.05) while results from distortion test suggested that the presence of horizontal pleiotropy had little impact on the most results (P>0.05) since there was no statistically significant difference in results after outlier removal. Besides, MR-Egger intercept analysis also suggested the absence of horizontal pleiotropy for most reported results (p>0.05) except BMI on coronary heart disease and BMI on hyperthyroidism.

## Discussion

This MR study found causal relationship between BMI and AF, and identified leptin, sleep apnoea and coronary heart disease as the mediators between BMI and AF. Specifically, around half of excess risk of AF related to obesity was mediated by circulating leptin. Furthermore, after adjustment for leptin and specific comorbidity (ie. sleep apnoea or coronary heart disease), the association between BMI and AF completely declined to null, indicating that targeting obesity-induced leptin resistance may be a promising strategy in preventing AF, especially for individuals with sleep apnoea or coronary heart disease.

Obesity has been well established as an important risk factor for AF.^3-8^ Both traditional observational epidemiological studies and MR studies have provided consistent evidence that obesity is associated with significant excess risk of AF. While the efficacy of weight-loss intervention in AF prevention was less well established, elucidating the underlying pathways between BMI and AF may contribute to providing a more precise strategy for AF prevention.^18,19^ We also found similar results in terms of the positive association between BMI and AF, what’s more, circulating leptin was identified as a dominating mediator in the BMI-AF association in present study.

Leptin, produced by adipose tissue, holds various regulatory functions in metabolism, immunity, and inflammation.^20^ It is reported that obesity leads to dramatic rise in circulating levels of leptin, and some experiments revealed that the reduction of leptin levels can impede the progression of obesity.^21^ Elevated leptin levels were demonstrated to be a cardiovascular risk factor.^22-24^ Some studies have reported the effect of leptin on AF but provided conflicting conclusions. For example, a case-control study found that individuals with AF had higher serum leptin levels compared to the control group.^25^ A MR study extensively explored the associations between visceral adipose tissue, circulating protein biomarkers and risk of cardiovascular diseases where a positive relationship between visceral fat, leptin levels and the risk of AF was observed.^26^ However, a prospective cohort study of postmenopausal women did not uncover a significant correlation between leptin or adiponectin and risk of AF.^27^ The exact mechanism by which leptin elevates the risk of AF remains unclear, although there were some hypotheses. For example, hyperleptinemia and leptin resistance, both associated with obesity, may contribute to the growth of left ventricular hypertrophy.^28^ A review reveals that dysfunction of the leptin signaling pathway is a significant contributor to obesity and related metabolic disorders and the effects of the leptin signaling pathway on the cardiovascular system are mediated through the modulation of cardiomyocyte and endothelial cell function, among other mechanisms.^29^ Animal experiments suggests that the signaling of leptin plays a role in the development of atrial fibrosis and the occurrence of AF triggered by angiotensin II or high-fat diet.^30,31^

It is surprising that the observed BMI-AF association completely declined to null in this study after adjustment for leptin and some comorbidity, such as sleep apnoea and atherosclerotic diseases (eg., coronary heart disease).

Obstructive sleep apnoea is a prevalent complication among individuals with obesity.^32,33^ A review has shown that obese individuals suffering from obstructive sleep apnoea have higher level of leptin compared to those without, indicating a positive relationship between obstructive sleep apnoea and leptin resistance.^34^ Additionally, previous research suggests that the disruption of leptin signaling in individuals with obstructive sleep apnoea may result in oxidative stress and increase the risk of cardiovascular diseases.^35^ A prospective analysis found that nearly half of AF patients suffered from sleep apnoea, while only 32% of the control group did.^36^ Some mechanisms behind the association between obstructive sleep apnoea and the increased risk of AF have been proposed, including recurrent nocturnal hypoxia/hypercapnia, surges in sympathetic tone and blood pressure during apneic episodes, and increased inflammation.^37,38^ These studies suggested interconnects between obesity, leptin, sleep apnoea and AF. This study found that the effect of obesity on AF was totally mediated by leptin and sleep apnoea, indicating that targeting leptin resistance induced by obesity and sleep apnoea may be an effective strategy for preventing AF.

Obesity is also associated with coronary heart disease, a risk factor for AF. Leptin secreted by adipose tissue may contribute to the development of atherosclerosis by affecting some types of cells, which may lead to the development of coronary heart disease.^24^ For example, previous studies suggested that leptin had the potential to enhance platelet aggregation, while which was only observed in obese individuals, suggesting the prothrombotic effect of leptin is probably restricted to individuals with obesity-induced leptin resistance and acts as a mediator between obesity and cardiovascular diseases.^10^ Similarly, this study found that the effect of obesity on AF was totally mediated by leptin and atherosclerotic diseases, suggesting that targeting obesity-induced leptin resistance may be effective in AF prevention through the inhibition of platelet aggregation among individuals at high risk of atherosclerotic diseases or with a history of atherosclerotic diseases. Besides, leptin may directly affect coronary endothelial cells by increasing the expression of tissue factor and cellular adhesion molecules.^39^

A major advantage of the study was that MR design could reduce the impact of confounding factors and avoid reverse causality as compared with traditional observational epidemiological studies, which can provide complementary evidence for the observational findings and examine the causal relationships between exposure, mediators and outcome. To our knowledge, this is the first MR study to extensively explore the potential mediating pathways between BMI and AF. However, there are several limitations of this study. First, the GWAS summary data used in this study are derived from relevant studies based on European populations, so the generalization of these findings is limited and further studies are needed in populations of other ethnicities. Second, individual-level GWAS data are not publicly available, we are thus not able to examine the associations in subgroup population. For example, previous study showed that there were sex differences in leptin levels, stratified analysis by sex may provide more information regarding the sex-specific mediating role of leptin. Third, sensitivity analyses showed that some instrumental variables may have horizontal pleiotropy, therefore the influence of potential horizontal pleiotropy on the results cannot be completely ruled out.

In conclusion, this MR study identified leptin as an important mediator between BMI and AF, suggesting that targeting leptin resistance may be crucial for developing effective prevention and treatment strategies to reduce the risk of obesity-related AF, particularly for individuals with sleep apnoea or coronary heart disease.

## Data Availability

All raw data are available in the public repository IEU open gwas project (https://gwas.mrcieu.ac.uk/)

## Abbreviations

AF: Atrial fibrillation
CRP: C-reactive protein
BMI: body mass index
MR: Mendelian randomization
GWASs: Genome-Wide Association Studies
LDL-C: low-density lipoprotein cholesterol
HDL-C: high-density lipoprotein cholesterol
TC: total cholesterol
TG: triglycerides
GIANT: Genetic Investigation of ANthropometric Traits
SNPs: single nucleotide polymorphisms
IVW-MR: inverse-variance weighted mendelian randomization
PERM: percentage of excess risk mediated
OR: odds ratio

## Contributors

All authors were responsible for the study concept and design. WH obtained funding. ZG did the statistical analysis and drafted the manuscript. WH and JX critically revised the manuscript. All authors gave the final approval and agreed to be accountable for all aspects of work ensuring integrity and accuracy.

## Acknowledgments

We thank the patients and investigators who contributed to the UK Biobank, GIANT Consortium, ADIPOGen, GLGC, International Consortium of Blood Pressure, FinnGen biobank, CARDIoGRAMplusC4D Consortium, ISGC Consortium.

